# Sociodemographic and health-related factors associated with viral load non-suppression and body mass index in adults with depression symptoms receiving antiretroviral therapy in South African primary care: secondary analysis of randomised trial data

**DOI:** 10.1101/2025.08.01.25332690

**Authors:** Babalwa Zani, Lara Fairall, Inge Petersen, Naomi Folb, Arvin Bhana, Graham Thornicroft, Jill Hanass-Hancock, Oné Selohilwe, Ruwayda Petrus, Sergio Carmona, Carl Lombard, Crick Lund, Naomi Levitt, Max Bachmann

## Abstract

**Introduction:** Unsuppressed viral load during antiretroviral therapy (ART) is associated with health decline and HIV transmission. Being overweight or obese increases the risk of non-communicable diseases, increasing the risk of multimorbidity in people living with HIV. Both ART effectiveness and obesity have been shown to be affected by socioeconomic, psychological and health related factors, but their interrelationships in South Africans living with HIV are not well known.

**Methods:** This was a secondary analysis of data from a randomised controlled trial of depression management in 2002 adults receiving ART. We investigated the effects of sociodemographic characteristics, comorbidities, depression symptoms (Patient Health Questionnaire-9 (PHQ-9)), functional disability (WHODAS-2.0), AIDS-related stigma and ART adherence on viral load non-suppression (viral load ≥1000 copies/ml) and on body mass index (BMI), at baseline (baseline) and on changes 12 months later, using longitudinal mixed effect logistic and linear regression models. Potentially confounding covariates were selected and adjusted for using least absolute shrinkage and selection operator (LASSO) inference.

**Results:** People with viral load non-suppression at baseline were more likely to be male, younger and to earn lower income. Health characteristics associated with viral load non-suppression at baseline were previous tuberculosis, having been on ART for less than 6 months or more than 10 years, and self-reported non-adherence to ART. Higher disability score and ART duration <6 months or >10 years at baseline were associated with an increasing likelihood of viral load non-suppression 12 months later. Higher BMI at baseline was associated with being female, being married, earning higher income and hypertension, no history of tuberculosis and not having viral load non-suppression. BMI increased from baseline to follow-up, and younger age was associated with a greater increase in BMI 12 months later. Depression symptom scores and stigma scores were not associated with viral load non-suppression or BMI.

**Conclusions:** Viral load non-suppression was associated with lower BMI, most likely due to its effects on HIV-related illness. Viral load non-suppression and BMI were both associated with a variety of sociodemographic factors, while viral load non-suppression was also associated with disability and ART non-adherence. These findings together indicate subgroups of people with HIV who most need improved ART access and adherence support. Neither outcome was associated with severity of depression symptoms or self-reported stigma.

ClinicalTrials.gov (NCT02407691), Pan African Clinical Trials Registry (201504001078347), South African National Clinical Trials Register (SANCTR) (DOH-27-0515-5048, NHREC 4048).

## Introduction

South Africa delivers the largest antiretroviral therapy (ART) programme globally, with over 5.7 million people receiving ART in 2022. The Joint United Nations Programme on HIV/AIDS (UNAIDS) estimated the prevalence of viral load suppression (HIV RNA<1000 copies/mL) to be at 92% in the treated population in South Africa in 2022 [1]. Viral load suppression rates in the treated population vary between provinces and districts. In the North West Province in 2015, at the time of this study, viral load suppression was estimated at 74% [2]. Viral load suppression is critical in HIV care as continued viral load non-suppression is associated with increased disease progression, health decline, acquisition of resistance to medication, higher risk of HIV transmission and death [3, 4]. Even in resource-constrained settings, viral load monitoring is key to achieving desired targets and identification of high-risk people for targeted interventions and factors which may compromise suppression need to be monitored.

A systematic review including studies from sub-Saharan Africa found that males, younger people and people who have lower household income were more likely to have viral load non-suppression [5]. Health characteristics associated with viral load non-suppression included sub-optimal adherence to ART, depression, having been on ART for a shorter duration, reporting a history of TB and having a higher BMI [5]. These studies reported the effect on viral load non-suppression of age, depression, duration on ART and history of TB among children and adolescents [6–11]. Most of these studies were cross-sectional, limiting inferences about direction of effect. Gaps also remain in the effects of ART history, health characteristics and disability on viral load non-suppression.

As ART programmes mature, PLWH are living longer and developing comorbidities, including cardiovascular and metabolic conditions. This can be attributed in part to the rising prevalence of overweight and obesity among PLWH. Obesity and higher BMI are increasing among PLWH as in the general population [12, 13]. Although weight gain after ART initiation may reduce mortality risk in underweight individuals, overweight and obesity increase the risk for high blood pressure as well as metabolic and cardiovascular disease [12, 14]. The prevalence of obesity is greater in Black women and in people of low socioeconomic status [12]. Excess caloric intake, high-fat diets, and reduced physical activity promote weight gain and obesity both among PLWH and in the general population [15, 16]. Studies from mostly the USA and Switzerland have shown high viral load to be associated with greater weight gain after ART initiation [17–19]. In South Africa, obesity among pregnant and post-partum women is more common with viral load non-suppression [20], but this relationship is unclear among the general population of PLWH.

We hypothesized that depression symptoms, stigma and disability would lead to lower adherence, which would lead to viral load non-suppression and worse clinical outcomes including weight loss. Comorbidities could also lead to increased depression symptoms and disability, with similar consequences for adherence and ART outcomes. We also hypothesized that viral load non-suppression, and adverse socioeconomic, psychological and physical health conditions, could be associated with BMI.

The aim of this study was to investigate the effects of depression symptom severity, stigma, disability, comorbidity and socioeconomic factors on viral load non-suppression and on BMI in people receiving ART who have depression symptoms. The specific objectives were to investigate whether these factors, measured at baseline, were associated with i) viral load non-suppression and BMI measured at baseline and ii) changes in viral load non-suppression and BMI from baseline to 12-month follow-up.

## Methods

### Study design and population

This study was a secondary analysis of data collected from participants enrolled in the Comorbid Affective Disorders, AIDS/HIV, and Long Term Health (CobALT) trial [21]. While the primary study used randomization to allocate participants to intervention and control arms, this secondary analysis study used data collected from participants in both arms combined. Data were collected at baseline and after 12 months follow-up. We investigated associations between two physical health outcomes – HIV viral load non-suppression and BMI – measured at baseline and at 12 months, and depression symptom severity, stigma, disability, comorbidity and socioeconomic factors measured at baseline.

CobALT was a pragmatic, two-arm cluster randomised trial conducted in 40 largest clinics in the Dr Kenneth Kaunda and Bojanala districts, North West province of South Africa. Participants were recruited from 21 April 2015 to 14 December 2016 [22] and follow-up interviews and data collection were conducted up to 18 January 2018. The trial assessed the effectiveness of a collaborative care model for the detection and management of depression on viral load non-suppression and depression symptoms among people on ART attending primary care facilities. Eligible participants were 18 years or older, on ART, and screened positive for depression symptoms as indicated by a score of nine or more on the PHQ-9, which was the established cut-off for depression in people with chronic conditions in the North West Province [23]. The trial showed that the intervention had no effects on depression symptoms or viral load non-suppression (the trial’s primary outcomes) or on any secondary outcomes including BMI [21].

### Data collection and management

We collected data using questionnaires administered by trained interviewers at baseline and at 12 months, separate from clinical care. Depression symptoms were assessed using a locally translated (Setswana) and validated version of the PHQ-9, with an inclusion cut-off score of nine [23, 24]. The PHQ-9 is a nine-item measure corresponding to the criteria upon which a diagnosis of depressive disorders is based in the Diagnostic and Statistical Manual of Mental Disorders (DSM-IV). Items are scored based on frequency of responses ranging from 0 (not at all) to 3 (nearly every day) and summed assuming equal weighting to yield a total score from 0 to 27, with higher scores indicating more severe depression symptoms.

The baseline questionnaire covered demographic details, level of education, employment status, income and self-reported comorbidities. Comorbidities were reported if a diagnosis had ever been given by a health care worker of stroke, heart attack or angina (ischaemic heart disease (IHD)), hypertension or tuberculosis (TB). The questionnaire included several standardised scales including the Internalised AIDS-Related Stigma Scale [25], the Perceived Stress Scale [26] and the 12-item World Health Organization Disability Assessment Schedule (WHODAS-2.0), which measures functional limitations [27–30]. We also recorded participants’ history on ART, including the date they initiated ART, the medication they were taking and the most recent viral load results. These interview data were complemented by examination of participants’ clinic folders. We used a visual analogue scale (VAS) ranging from 0-100% to measure self-reported adherence to ART in the last month as reported to trained fieldworkers separate from clinical care. Values below 90% were categorised as non-adherent [31]. Duration on ART was calculated as the number of months between the time the participant-initiated ART and the date of their baseline interview.

We measured weight using a digital scale and height using a wall mounted height meter and used these to calculate BMI, defined as the body mass divided by the square of the body height (kg/m^2^). We categorised ART duration as < 6 months, 6 to 120 months, and > 120 months. Longer duration on ART was categorised as having been on ART for 10 years (120 months) or more, as a previous study in South Africa showed an increase in the probability of virological failure and of switching to second-line therapy after being on ART for 10 years [32].

Viral load was assessed at baseline and at 12 months. At baseline in the Dr Kenneth Kaunda district, we recorded the most recent available viral load result from clinic files. For study participants with no viral load in the preceding six months, we collected blood samples for viral load analysis. Follow-up viral loads were assessed for all participants. In the Bojanala district, we collected blood samples for all participants at baseline and follow-up. We also obtained routine viral load results from the National Laboratory Health Services (NHLS). Blood samples for research-funded viral load analysis were collected at the clinics using standard procedures and processed with routine blood samples at the NHLS.

We defined viral load non-suppression as HIV RNA ≥1000 copies/mL [33]. Participants with missing viral load results were excluded in the analysis.

### Statistical analysis

First, we investigated crude associations between participants’ depression symptom severity, stigma and disability scores, comorbidities, socioeconomic factors (the explanatory or independent variables) and viral load non-suppression and BMI (outcome variables) measured at baseline and 12-month follow-up. PHQ-9 score, stigma score and disability score were standardised by subtracting the mean from each measurement and dividing by the standard deviation for use as regression covariates. P-values were estimated for each explanatory variable separately, using logistic regression models (with viral load non-suppression as outcome) or linear regression models (with BMI as outcome), with robust adjustment for clustering of outcomes within clinics [34, 35]. To compare the magnitude of differences between participants with and without detectable viral loads among variables measured with different scales, we calculated standardized differences in baseline variables using Cohen’s formulae for differences in means and proportions [36].

Secondly, we investigated the mutually adjusted associations between the explanatory variables, measured at baseline, and the viral load non-suppression and BMI outcomes measured at baseline and 12-month follow-up, using multilevel longitudinal mixed effects regression models [37]. A linear model was used for BMI as a continuous outcome and a generalized linear model with a logit link was used for viral load non-suppression as binary outcome. Individuals and clinics were modelled as having random intercepts, with individuals nested within clinics, to account for repeated measurement of outcomes in individuals, and randomisation of clinics. The explanatory variables of interest, recorded at baseline, were sociodemographic indicators (age, sex, marital status, household income); depression symptoms (PHQ-9), stigma and disability scores; comorbid conditions (hypertension, IHD, previous TB) and ART history (non-adherence, ART started less than 6 months or more than 10 years ago). For models with BMI as an outcome we used viral load non-suppression instead of ART history as an explanatory variable because we assumed that effects of ART history on BMI would be mediated through viral load non-suppression. All models included trial arm (intervention or control) as a covariate to account for the origin of the data in a randomised trial, although the purpose of present study was not to estimate the effects of the intervention.

To account for the longitudinal study design, data were arranged so that each participant had two records. The first record comprised baseline measures of the explanatory variables, the baseline measures of viral load non-suppression and BMI, trial arm, and a dummy time variable coded as 0. The second record comprised the same baseline measures of the explanatory variables, the 12-month follow-up measures of viral load non-suppression and BMI, and a dummy time variable coded as 1. To estimate the effects of each baseline explanatory variable on changes in outcomes we included covariate-time interaction variables. The coefficients for these interaction variables quantified the estimated effect of the baseline explanatory variables on changes in the outcome from baseline to 12-month follow-up.

Thirdly, to investigate whether these models may have missed possible statistically significant associations between outcomes and explanatory variables because of over-adjustment for numerous correlated covariates, we constructed more parsimonious regression models using LASSO (least absolute shrinkage and selection operator) inference methods [38, 39]. For each explanatory variable of interest, LASSO algorithms select a subset of the covariates that are associated either with the outcome of interest, or with the explanatory variable of interest, or both. Covariate selection is based on the magnitude of their coefficients and not on p-values. The selected variables are then included in a final linear or logistic regression model which estimates the adjusted association between the explanatory variable of interest and the outcome. We used Stata’s double selection inference applications ‘dsregress’ and ‘dslogit’ [40]. Because these applications do not support multilevel mixed models, we used separate models for outcomes measured at baseline and at follow-up, with variance correction for clustering of outcomes within clinics. Models with each 12-month follow-up value as outcome included baseline values of the same variable as a covariate; the model coefficients therefore represent associations between the explanatory variables and changes in the outcome [41].

Statistical analyses were performed with Stata version 18 statistical software [42]. A P-value of 0.05 or less was considered statistically significant.

### Ethical considerations and trial registration

The trial was registered with ClinicalTrials.gov (NCT02407691), the Pan African Clinical Trials Registry (201504001078347) and the South African National Clinical Trials Register (DOH-27-0515-5048NHREC number 4048). Ethical approval for the trial was obtained from the University of Cape Town Human Research Ethics Committee (211/2013), King’s College London Research Ethics Office (PNM/12/13-159) and the University of KwaZulu-Natal Biomedical Research Ethics Committee (BFC049/15). Permission to conduct the study was granted by the North West Provincial Department of Health. All patient participants provided written informed consent to participate in the study. The study was monitored by a Data and Safety Monitoring Board (DSMB), holding meetings twice a year for the duration of the study.

### Results

A total of 2002 participants were enrolled and interviewed at baseline, 1008 in the intervention and 994 in the control arm; of whom 1675 (84%) were interviewed again at 12 months. Of the enrolled participants, 1876 (94%) had viral load results available at baseline and 1837 (92%) at 12 months. Socioeconomic, health and clinic characteristics of the participants are shown in Table 1. Most participants were black, women and unmarried, and half were aged 42 years or older. Just under three-quarters were unemployed, with over half receiving a government social grant. The average personal monthly income was low: equivalent to US$ 72.05 in 2016 (ZAR 1060) [43]. The mean PHQ-9 score at baseline was 14, with 65% of participants having a score of 9-14 corresponding to mild depression symptoms.

**Table 1:** Socioeconomic, psychological and health characteristics of participants with or without viral load suppression at baseline and at 12 months

| Characteristics | At baseline |  |  |  |  | At 12 months |  |  |  |  |
| --- | --- | --- | --- | --- | --- | --- | --- | --- | --- | --- |
|  | All participants<sup>1</sup><br>n (%) | Viral load ≥1000<br>copies/ml: n (%) | Viral load <1000<br>copies/ml: n (%) | sMD | p-value* | All participants<sup>1</sup><br>n (%) | Viral load ≥1000<br>copies/ml: n (%) | Viral load <1000<br>copies/ml: n (%) | sMD | p-value* |
| Total participants | N=1876 | 1574 (86%) | 302 (914%) |  |  | N=1837 | 1585 (85%) | 282 (15%) |  |  |
| Socioeconomic characteristics |  |  |  |  |  |  |  |  |  |  |
| Age (years): mean (SD) | 42(11) | 40 (10) | 42 (11) | 0.20 | 0.004 | 42 (11) | 40 (11) | 43 (11) | 0.23 | 0.002 |
| Male | 340 (18) | 78 (23) | 262 (77) | 0.23 | <0.001 | 320 (17) | 65 (20) | 255 (80) | 0.17 | 0.023 |
| Female | 1536 (82) | 224 (15) | 1312 (85) |  |  | 1517 (83) | 217 (14) | 1300 (86) |  |  |
| Married | 437 (23) | 55 (13) | 382 (87) | 0.15 | 0.034 | 415 (23) | 51 (12) | 364 (88) | 0.13 | 0.067 |
| Unmarried | 1439 (77) | 247 (17) | 1192 (83) |  |  | 1422 (77) | 231 (16) | 1191 (84) |  |  |
| Total income <sup>2, 3</sup> | 1.1 (1.7) | 0.8 (1.5) | 1.1 (1.8) | 0.15 | 0.042 |  | 0.9 (1.63) | 1.1 (1.8) | 0.11 | 0.172 |
| Psychological characteristics, disability and BMI |  |  |  |  |  |  |  |  |  |  |
| PHQ-score: mean (SD) | 13.7 (4.0) | 14.2 (4.1) | 13.6 (4.0) | -0.13 | 0.023 | 13.7 (4.0) | 13.9 (4.2) | 13.7 (4.0) | -0.05 | 0.431 |
| Stress score: mean (SD) | 20.8 (6.6) | 20.9 (6.2) | 20.7 (6.7) | -0.02 | 0.770 | 20.7 (6.6) | 21.1 (6.3) | 20.6 (20.3) | -0.07 | 0.319 |
| Stigma score: mean (SD) | 2.7 (2.1) | 2.5 (2.1) | 2.8 (2.1) | 0.12 | 0.082 | 2.7 (2.1) | 2.5 (2.3) | 2.7 (2.6) | 0.11 | 0.098 |
| Disability score: mean (SD) | 10.4 (7.7) | 10.7 (7.5) | 10.3 (7.9) | -0.06 | 0.345 | 10.3 (7.7) | 11.2 (7.6) | 10.1 (7.7) | -0.14 | 0.053 |
| BMI | N=1854 |  |  |  |  | N=1812 |  |  |  |  |
| Mean (SD) | 26.5 (7.9) | 22.4 (5.5) | 25.0 (6.8) | 0.38 | <0.001 | 25.0 (6.7) | 22.9 (5.8) | 25.0 (6.8) | 0.31 | 0.008 |
| ART experience |  |  |  |  |  |  |  |  |  |  |
| Self-reported adherence <90% at baseline | 211 (11) | 67 (32) | 144 (68) | 0.36 | <0.001 | 193 (11) | 59 (31) | 134 (69) | 0.35 | <0.001 |
| Self-reported adherence ≥90% at baseline | 1665 (89) | 235 (14) | 1430 (86) |  |  | 1644 (89) | 223 (14) | 1421 (86) |  |  |
| ART duration 6 months to 10 years | 1347/1614 (83) | 167 (12) | 1180 (88) | 0.32 | <0.001 | 1290/1564 (82) | 168 (13) | 1122 (87) | 0.14 | 0.028 |
| ART duration < 6 months or > 10 years | 267/1614 (17) | 65 (24) | 202 (76) |  |  | 274/1564 (18) | 52 (19) | 222 (81) |  |  |
| Viral load non-suppression at baseline |  |  |  |  |  | 274/1724 (16) | 160 (58) | 114 (42) | -1.72 | <0.001 |
| Viral load suppression at baseline |  |  |  |  |  | 1450/1724 (84) | 101 (7) | 1349 (93) |  |  |
| Self-reported comorbidity |  |  |  |  |  |  |  |  |  |  |
| Hypertension diagnosis or treatment | 591 (32) | 90 (15) | 501 (85) | 0.04 | 0.461 | 589 (32) | 79 (13) | 510 (87) | 0.10 | 0.181 |
| No hypertension diagnosis or treatment | 1285 (68) | 212 (16) | 1073 (84) |  |  | 1248 (68) | 203 (16) | 1045 (84) |  |  |
| Ischaemic heart disease | 404/1874 (22) | 82 (20) | 322 (80) | 0.05 | 0.009 | 411 | 68 (17) | 343 (83) | 0.03 | 0.446 |
| No ischaemic heart disease | 1470/1874 (78) | 220 (15) | 1250 (85) |  |  | 1423 | 214 (15) | 1209 (845) |  |  |
| Previous tuberculosis diagnosis | 660/1874 (35) | 147 (22) | 513 (78) | 0.33 | <0.001 | 659/1834 (36) | 129 (20) | 530 (80) | 0.24 | <0.001 |
| No previous tuberculosis diagnosis | 1214/1874 (65) | 155 (13) | 1059 (87) |  |  | 1175/1834 (64) | 153 (13) | 1022 (87) |  |  |

<sup>1</sup>The denominator for variables that have missing responses are indicated individually; for other variables the denominators are total participants.<sup>2</sup>Total income is total grant, employment and business income per person.
<sup>3</sup>1000 South African Rand per month per person. Smd= Standardised mean difference. SD=standard deviation. \*P values from logistic regression models with robust adjustment for intra-clinic correlation of outcomes

Of 1876 participants with viral load results, 1574 (84%) had viral load suppression at baseline and 1585 (85%) at 12 months (Table 1). Viral load suppression varied across sub-districts, ranging from 75% to 94% baseline and 76% to 93% at 12 months. At baseline, 11% of participants reported ART adherence of less than 90%. The median duration on ART at baseline was 37.9 months (interquartile range (IQR) 14.9-65.7 months); 12% had been on ART for less than 6 months and 5% for more than 120 months.

### Factors associated with viral load non-suppression at baseline and 12 months

In the bivariable analysis (Table 1), viral load non-suppression at baseline was significantly more common in men, in participants who were younger, unmarried, and those who earn a lower income. Participants with viral load non-suppression had higher mean scores for depressive symptoms, lower mean BMI, more frequently reported previous IHD, previous TB diagnosis, lower levels of ART adherence and having been on ART for less than 6 months or more than 120 months. Unsuppressed viral load at 12 months followed similar patterns, however, depressive symptoms, income and IHD no longer showed a significant difference (Table 1). Standardized differences between participants with and without viral load suppression were greatest for BMI, nonadherence to ART, tuberculosis and duration of ART at baseline and for BMI and nonadherence to ART at 12 months.

In the mixed effects logistic regression model (Table 2), socioeconomic characteristics associated with viral load non-suppression at baseline were younger age, being male, and earning a lower income. Health characteristics associated with viral load non-suppression were self-reported non-adherence to ART, having been on ART for less than 6 months or more than 10 years (120 months) and reporting a history of tuberculosis. Higher disability score at baseline was associated with increasing likelihood of having viral load non-suppression from baseline to follow-up and having been on ART for less than 6 months or more than 10 years at baseline was associated with deceasing likelihood of viral load non-suppression from baseline to follow-up. The LASSO models showed similar associations, except that the association between ART duration less than 6 months or more than 10 years was not associated with reduced odds of having viral load non-suppression at 12 months (Table 3).

**Table 2:** Sociodemographic and health characteristics measured at baseline and their association with viral load non-suppression at baseline, and with changes in viral load non-suppression from baseline to 12 months: longitudinal logistic regression mixed model including all covariates

| Characteristics | Outcome: Viral load non-suppression (HIV RNA $\geq 1000$ copies/ml) | | |
| --- | --- | --- | --- |
|  | OR | 95% CI | p-value |
| **Outcome: Viral load non-suppression (HIV RNA $\geq 1000$ copies/ml) at baseline** |  |  |  |
| **Socioeconomic characteristics** |  |  |  |
| Age (per year) | 0.96 | 0.93; 0.99 | **0.005** |
| Female | 0.29 | 0.14; 0.61 | **0.001** |
| Married | 0.58 | 0.27; 1.20 | 0.142 |
| Total income (per ZAR 1000/month) | 0.80 | 0.64; 0.98 | **0.032** |
| **Psychological and health characteristics** |  |  |  |
| PHQ-9 score (per standard deviation) | 1.16 | 0.85; 1.59 | 0.341 |
| Stigma score (per standard deviation) | 0.91 | 0.68; 1.22 | 0.519 |
| Disability score (per standard deviation) | 1.05 | 0.76; 1.45 | 0.779 |
| Ischaemic heart disease | 1.69 | 0.72; 3.97 | 0.224 |
| Hypertension diagnosis | 1.23 | 0.63; 2.41 | 0.551 |
| History of TB | 3.01 | 1.61; 5.65 | **0.001** |
| **ART experience** |  |  |  |
| Self-reported adherence $\geq 90\%$ | 0.10 | 0.04; 0.23 | **<0.001** |
| ART duration <6 months or >120 months. vs 6-120 months | 8.17 | 3.57; 18.70 | **<0.001** |
| **Trial design** |  |  |  |
| Intervention versus control arm | 0.90 | 0.41; 1.99 | 0.788 |
| **Outcome: Change in viral load non-suppression (HIV RNA $\geq 1000$ copies/ml) at 12 months<sup>1</sup>** |  |  |  |
| **Socioeconomic characteristics** |  |  |  |
| 12-months vs. baseline | 0.74 | 0.14; 3.66 | 0.699 |
| Age | 1.01 | 0.98; 1.04 | 0.585 |
| Female | 1.44 | 0.68; 3.05 | 0.347 |
| Marriage | 1.23 | 0.57; 2.65 | 0.596 |
| Income | 1.04 | 0.84; 1.28 | 0.751 |
| **Psychological and health characteristics** |  |  |  |
| PHQ-9 score (per standard deviation) | 0.83 | 0.61; 1.13 | 0.234 |
| Stigma score (per standard deviation) | 0.89 | 0.66; 1.20 | 0.459 |
| Disability score (per standard deviation) | 1.50 | 1.08; 2.08 | **0.017** |
| Ischaemic heart disease | 0.62 | 0.26; 1.47 | 0.276 |
| Hypertension diagnosis | 0.62 | 0.31; 1.24 | 0.173 |
| History of TB | 0.62 | 0.33; 1.16 | 0.131 |
| **ART experience** |  |  |  |
| Self-reported adherence $\geq 90\%$ | 1.32 | 0.58; 2.98 | 0.503 |
| ART duration <6 months or >120 months. vs 6-120 months | 0.30 | 0.13; 0.69 | **0.004** |
| **Trial design** |  |  |  |
| Intervention versus control arm | 0.79 | 0.44; 1.43 | 0.443 |

<sup>1</sup>ORs for change in outcome estimated with covariate time interaction terms. OR=Odds Ratio, CI=Confidence Interval. Covariates retained in model if adjusted P<0.01.

**Table 3:** Risk factors associated with viral load non-suppression at baseline and with changes in viral load non-suppression from baseline to 12 months: logistic regression models with odds ratio for each variable adjusted for potentially confounding covariates selected using LASSO

| Outcome and explanatory variables | Viral load non-suppression (HIV RNA $\geq 1000$ copies/ml) | | | |
| --- | --- | --- | --- | --- |
|  | OR | 95% CI | P-value | selected control variables |
| **Outcome: Viral load non-suppression (HIV RNA $\geq 1000$ copies/ml) at baseline<sup>1</sup>** |  |  |  |  |
| **Socioeconomic characteristics** |  |  |  |  |
| Age (per year) | 0.98 | 0.96; 0.99 | **0.002** | sex, hypertension, ischaemic heart disease, |
| Female | 0.63 | 0.44; 0.90 | **0.011** | age |
| Married | 0.74 | 0.51; 1.08 | 0.119 | - |
| Total income (per ZAR 1000/month) | 0.88 | 0.77; 1.01 | 0.073 | age |
| **Psychological and health characteristics** |  |  |  |  |
| PHQ-9 score (per standard deviation) | 1.11 | 0.97; 1.27 | 0.138 | disability |
| Stigma score (per standard deviation) | 0.94 | 0.81; 1.08 | 0.353 | disability |
| Disability score (per standard deviation) | 0.99 | 0.86; 1.14 | 0.890 | stigma, hypertension, ischaemic heart disease |
| Ischaemic heart disease | 1.33 | 0.94; 1.89 | 0.104 | age, depression, hypertension |
| Hypertension diagnosis | 1.16 | 0.87; 1.53 | 0.306 | age, disability, ischaemic heart disease |
| History of TB | 1.88 | 1.43; 2.47 | **<0.001** | sex, disability, ischaemic heart diseases |
| Self-reported adherence $\geq 90\%$ | 0.35 | 0.24; 0.49 | **<0.001** | - |
| ART duration <6 months or >120 months (vs 6-120 months) | 2.74 | 1.90; 3.94 | **<0.001** | age, adherence |
| **Outcome: Change in viral load non-suppression (HIV RNA $\geq 1000$ copies/ml) from baseline to 12 months<sup>2</sup>** |  |  |  |  |
| **Socioeconomic characteristics** |  |  |  |  |
| Age (per year) | 0.99 | 0.97; 1.01 | 0.242 | sex, hypertension, ischaemic heart disease |
| Female | 0.95 | 0.56; 1.60 | 0.836 | age |
| Married | 0.93 | 0.57; 1.54 | 0.787 | depression |
| Total income (per ZAR 1000/month) | 0.94 | 0.81; 1.08 | 0.356 | - |
| **Psychological and health characteristics** |  |  |  |  |
| PHQ-9 score (per standard deviation) | 0.92 | 0.75; 1.12 | 0.385 | - |
| Stigma score (per standard deviation) | 0.85 | 0.71; 1.02 | 0.083 | - |
| Disability score (per standard deviation) | 1.35 | 1.12; 1.62 | **0.001** | depression, stigma, ischaemic heart disease |
| Ischaemic heart disease | 0.88 | 0.57; 1.34 | 0.541 | age, disability |
| Hypertension diagnosis | 0.80 | 0.50; 1.27 | 0.338 | age |
| History of TB | 1.09 | 0.76; 1.55 | 0.650 | sex |
| Self-reported adherence $\geq 90\%$ | 0.67 | 0.39; 1.16 | 0.149 | - |
| ART duration <6 months or >120 months (vs 6-120 months) | 0.73 | 0.15; 0.43 | 0.245 | income |

<sup>1</sup>adherence, art duration and tuberculosis also included as control variables in all models; <sup>2</sup>associations with change in viral load non- suppression estimated by including viral load non-suppression at baseline as covariate in all models, disability also included as control variable in all models

### Factors associated with body mass index

At baseline, 1970 (98%) participants had BMI values recorded and 1562/1675 (93%) had BMI values recorded at 12 months. Mean BMI was 24.6 (standard deviation (SD) 6.7) kg/m^2^ at baseline and 24.9 (SD 6.9) kg/m^2^ at 12 months. In the mixed effect linear regression model, being female, married, earning a higher income, and having hypertension were associated with greater BMI at baseline, and history of tuberculosis and viral load non-suppression were associated with lower BMI at baseline (Table 4). In the same model, BMI increased from baseline to follow-up, and older age was associated with a lesser increase from baseline to follow-up while being female was associated with a greater increase. The LASSO models showed similar associations, except that ischaemic heart disease was also associated with increasing BMI from baseline to follow-up (Table 5).

**Table 4:** Association of socioeconomic, psychological and health characteristics measured at baseline with BMI at baseline and with changes in BMI from baseline to 12 months: longitudinal linear regression mixed model including all covariates

| Characteristics | BMI |  |  |
| --- | --- | --- | --- |
|  | Coefficient | 95% CI | p-value |
| **Outcome: BMI at baseline** |  |  |  |
| **Socioeconomic characteristics** |  |  |  |
| Age (per year) | 0.01 | -0.02; 0.03 | 0.731 |
| Female | 4.91 | 4.16; 5.66 | **0.001** |
| Married | 0.76 | 0.09; 1.42 | **0.026** |
| Total income (per ZAR 1000/month) | 0.30 | 0.14; 0.46 | **0.001** |
| **Psychological and health characteristics** |  |  |  |
| PHQ-9 score (per standard deviation) | -0.13 | -0.44; 0.17 | 0.395 |
| Stigma score (per standard deviation) | 0.06 | -0.23; 0.34 | 0.694 |
| Disability score (per standard deviation) | -0.17 | -0.48; 0.14 | 0.277 |
| Ischaemic heart disease | -0.05 | -0.88; 0.78 | 0.909 |
| Hypertension diagnosis | 2.26 | 1.61; 2.90 | **0.001** |
| History of TB | -1.45 | -2.04; -0.85 | **0.001** |
| Viral load non-suppression at baseline | -0.54 | -0.92; -0.15 | **0.007** |
| **Trial design** |  |  |  |
| Intervention versus control arm | -0.15 | -1.05; 0.75 | 0.737 |
| **Outcome: BMI at 12 months** |  |  |  |
| **Socioeconomic characteristics** |  |  |  |
| Time from baseline to 12-months | 0.80 | 0.13; 1.47 | **0.019** |
| Age (per year) | -0.02 | -0.04; -0.01 | **0.001** |
| Female | 0.38 | 0.04; 0.73 | **0.029** |
| Married | -0.18 | -0.48; 0.12 | 0.238 |
| Total income (per ZAR 1000/month) | -0.002 | -0.08; 0.08 | 0.970 |
| **Psychological and health characteristics** |  |  |  |
| PHQ-9 score (per standard deviation) | -0.07 | -0.20; 0.07 | 0.346 |
| Stigma score (per standard deviation) | -0.04 | -0.17; 0.09 | 0.520 |
| Disability score (per standard deviation) | 0.09 | -0.05; 0.23 | 0.198 |
| Ischaemic heart disease | -0.35 | -0.71; 0.00 | 0.053 |
| Hypertension diagnosis | 0.02 | -0.27; 0.30 | 0.908 |
| History of TB | 0.07 | -0.20; 0.34 | 0.614 |
| Viral load non-suppression at baseline | 0.23 | -0.19; 0.64 | 0.286 |
| **Trial design** |  |  |  |
| Intervention versus control arm | 0.14 | -0.11; 0.39 | 0.279 |

**Table 5:** Factors associated with BMI at baseline and with change in BMI from baseline to 12 months: linear regression models with coefficients for each variable adjusted for potentially confounding covariates selected by LASSO

| Outcome and explanatory variables | BMI |  |  |  |  |
| --- | --- | --- | --- | --- | --- |
|  | coefficient | 95% CI |  | P-value | selected control variables |
| Outcome: BMI at baseline<sup>1</sup> |  |  |  |  |  |
| Age (per year) | 0.01 | -0.02 | 0.05 | 0.494 | ischaemic heart disease |
| Female | 4.89 | 4.27 | 5.51 | <0.001 | age, income |
| Married | 1.03 | 0.34 | 1.71 | 0.003 |  |
| Total income (per ZAR 1000/month) | 0.29 | 0.11 | 0.47 | 0.001 | age, income |
| PHQ-9 score (per standard deviation) | -0.21 | -0.51 | 0.10 | 0.179 | disability |
| Stigma score (per standard deviation) | 0.13 | -0.17 | 0.43 | 0.393 | disability |
| Disability score (per standard deviation) | -0.21 | -0.56 | 0.15 | 0.250 | depression, stigma, ischaemic heart disease |
| Ischaemic heart disease | -0.04 | -0.89 | 0.81 | 0.921 | age, depression |
| Hypertension diagnosis | 2.11 | 1.23 | 2.98 | <0.001 | age |
| History of TB | -1.36 | -1.98 | -0.74 | <0.001 | disability, ischaemic heart disease |
| Viral load non-suppression at baseline | -1.79 | -2.42 | -1.15 | <0.001 |  |
| Outcome: Change in BMI at 12 months<sup>2</sup> |  |  |  |  |  |
| Age (per year) | -0.02 | -0.03 | -0.01 | <0.001 | hypertension |
| Female | 0.65 | 0.25 | 1.06 | 0.001 | age |
| Married | -0.14 | -0.46 | 0.18 | 0.399 |  |
| Total income (per ZAR 1000/month) | 0.03 | -0.04 | 0.09 | 0.413 | age |
| PHQ-9 score (per standard deviation) | -0.09 | -0.25 | 0.06 | 0.239 | disability |
| Stigma score (per standard deviation) | -0.03 | -0.15 | 0.09 | 0.596 | disability |
| Disability score (per standard deviation) | 0.04 | -0.07 | 0.15 | 0.509 | depression, stigma, ischaemic heart disease |
| Ischaemic heart disease | -0.40 | -0.67 | -0.13 | 0.004 | depression, hypertension, tuberculosis |
| Hypertension diagnosis | 0.13 | -0.21 | 0.48 | 0.452 | age, ischaemic heart disease |
| History of TB | -0.05 | -0.34 | 0.23 | 0.711 |  |
| Viral load non-suppression at baseline | 0.19 | -0.18 | 0.55 | 0.319 | tuberculosis |

<sup>1</sup>All models include sex, hypertension, tuberculosis and viral load non-suppression at baseline as control variables; <sup>2</sup> Associations with change in BMI estimated by including BMI at baseline as covariate in all models; sex also included as control variable in all models

## Discussion

This study showed that sociodemographic characteristics associated with viral load non-suppression among PLWH with depressive symptoms, reported in the present study, were similar to those reported in previous studies among the general adult population of PLWH and among adolescents. The present study showed, in addition, that very short or very long exposure to ART was associated with having viral load non-suppression, and that disability was associated with increased risk of viral load non-suppression over time. This study also confirmed the characteristics associated with higher BMI and with increasing BMI over time.

Previous studies in South Africa have shown functional disability to be associated with reduced adherence to ART [28, 44]. This study further highlights its longer term role in viral load non-suppression, which are likely to be due to greater difficulty accessing and adhering to ART. Disability scores among people with depressive symptoms in this study were higher than previously reported among PLWH in South Africa [28]. Higher disability scores are an indication of functional limitations and if not addressed, may pose a risk of physical disability, lower livelihood outcomes and increased depression symptoms [29]. ART programmes need to consider disability, mental health and rehabilitation services as part of their care package.

Although depressive symptoms at ART initiation were associated with viral load non-suppression [45–47], in this study the severity of depressive symptoms as well as of stigma had no effect on either viral load non-suppression or BMI.

The study included people who had been on ART for varying times. ART significantly reduces HIV viral load within the first few days, with many people having undetectable viral load within 1 – 3 months [48]. However, the reduction in viral load may be affected by adherence. Previous research has shown that people who have been on ART for longer durations may experience treatment fatigue due to pill burden and patient-provider interactions [49]. It is thus critical to continue monitoring viral load and to provide long-term psychological support to people on ART. While self-reported ART adherence measured using VAS is subjective and prone to over-estimation, it has been shown to be strongly associated with other measures of adherence [50]. Self-reported adherence has shown reasonable correlation with viral load and other objective markers of adherence even in low-resourced settings and is more trustworthy when collected separately from clinical care [51–53]. Our finding that self-reported ART adherence was associated with viral load non-suppression is therefore unsurprising.

Findings of higher BMI among women compared to men, and its association with hypertension are consistent with previous studies, highlighting a population at risk of this cluster of multimorbidity [17, 54, 55]. The rise in obesity among PLWH, especially women, may simply reflect the increasing rate of obesity in the general population. This may be influenced by South Africa’s transition to urbanization and more energy dense dietary patterns. The association of higher BMI with higher income, although in a generally impoverished population, was consistent with a previous study [56] and is likely reflective of increased access and consumption of high fat and sugar diets and access to fast foods. Cultural perceptions of a ‘healthy’ weight, where lower weight is perceived as a sign of ill-health or an indication of living with HIV, and higher weight perceived as a sign of better health, greater prosperity and being more common among people without HIV [56–59] may be contributing to obesity through excess caloric intake and consumption of high-fat diets.

The findings that both male sex and younger age are associated with viral load non-suppression is concerning. Both behavioural and structural factors may be contributing to a significantly higher risk of having viral load non-suppression in men with HIV and depression symptoms. Men tend to present to health facilities with advanced disease [60–62] and engage in alcohol consumption in the evenings which may make it more difficult to take ART [63]. There have been concerns as to whether primary care clinics in their current format are able to attract young people and men or whether alternative modes of care are required [64, 65]. It is therefore important to have interventions that focus on men and young people who are on ART and at risk of depression.

This study had several limitations. Firstly, the high prevalence of viral load suppression in this clinic-based population constrained our statistical power to identify risk factors for viral load non-suppression. A community sample including those who have disengaged with care may have provided more insight. Secondly, we did not assess alcohol and substance misuse, which have been associated with depression, especially in men and people disengaged from care [66, 67], and with virological failure [68]. This decision was informed by a low case-detection rate for alcohol misuse disorder in our pilot study in the same population [69].

Thirdly, we did not record or adjust for ART regimen although second and third line regimens might be associated with virological failure. However, 94% of the participants were known to be on the first-line regimen. Fourthly, as CD4 counts were no longer routinely measured, we did not assess it although it is an important indicator of immunological status and of adherence. We did not assess drug resistance, which has been reported to be at 10% in low- and middle-income countries [70]. Fifthly, although weight loss is a hallmark of HIV/AIDS, lower BMI can be an ambiguous outcome in settings such as this in which overweight and obesity are highly prevalent. Studies have also associated weight gain with ART initiation [17, 55], however, our study did not explore the different regimens that could be associated with higher BMI. Sixthly, we also did not compare depressive symptoms among people receiving ART and those not living with HIV. Seventhly, the 12-month follow-up period limited our analysis of changes in outcomes. Finally, this study was conducted in South Africa, among a generally impoverished population in a country with crippling inequality [71]. A country also facing a crisis of obesity among the adult population [72]. The applicability of these findings in other contexts may differ.

The strengths of the study included the use of longitudinal data, enabling us to identify characteristics associated with viral load non-suppression and BMI at baseline and with changes during follow-up. We also used objective measures of outcomes, including our viral load data which included three sources which were triangulated. The findings of multilevel longitudinal mixed effect regression models were confirmed by the parsimonious regression models using LASSO inference methods.

## Conclusions

In conclusion, this study identifies subgroups of PLWH and on ART who may benefit from targeted interventions for viral load and weight management, including improving access and supporting adherence. The study also identifies the long-term effects of functional disability on viral load. Viral load monitoring and support should be consistent and be targeted at those who have recently initiated ART or may be experiencing treatment fatigue. The management of HIV should encompass accurate information on weight management and prevention of multimorbidity.

## Authors’ contributions

LF, IP, GT, NF, AB, CLom, MB, CLun, NL, JHH, BZ and SC participated in designing the study; LF, IP, GT, NF, AB, CLom, MB, CLun, NL and JHH participated in applying for funding; IP, LF, LF, NF, IP, AB, GT, OS and BZ participated in planning the data collection; BZ managed field data collection; BZ and MB statistically analysed the data; BZ wrote the first draft; all authors provided comments and approved the final manuscript.

## Data Availability

All MS Excel files are available from the Open Science Framework database

## Acknowledgements

The authors thank the clinic managers, phlebotomists and data capturers who assisted with study activities at the clinics. We thank Mrs. Deanna Carter and Daniella Georgeu-Pepper for supporting training of fieldworkers and fieldwork activities. We thank the fieldworkers who collected the data and assisted with tracing study participants for follow-up and the people who participated. We thank the staff at the NHLS for assisting with analysis of viral load samples and provision of results.

## Funding and disclaimers

The research reported in this publication was supported by the National Institute of Mental Health of the National Institutes of Health under Award Number R01MH100470. The content is solely the responsibility of the authors and does not necessarily represent the official views of the National Institutes of Health. This study is also an output of the Programme for improving mental health care (PRIME) and was supported by the UK Department for International Development [201446]. The views expressed do not necessarily reflect the UK Government’s official policies. BZ received partial funding by the South African Medical Research Council through the Division of Research Capacity Development under the Researcher Development Grant. The contents thereof are the sole responsibility of the authors and does not necessarily represent the official views of the SAMRC.

